# SARS-CoV-2 Immunogenicity in individuals infected before and after COVID-19 vaccination: Israel, January-March 2021: Implications for vaccination policy

**DOI:** 10.1101/2021.04.11.21255273

**Authors:** Kamal Abu Jabal, Hila Ben-Amram, Karine Beiruti, Ira Brimat, Ashraf Abu Saada, Younes Bathish, Christian Sussan, Salman Zarka, Michael Edelstein

## Abstract

Between December 2020-March 2021 we measured anti-SARS-CoV-2 IgG titers post-vaccination with the BNT162b2 vaccine among 725 Israeli hospital workers. Previously infected individuals who received one dose had higher IgG titres than fully vaccinated, never-infected workers. Post-vaccination infection did not increase IgG titres. Individuals infected post-dose one should receive the second.

## Background

As of March 2021, Israel was the country with the highest COVID19 vaccine coverage1). It used the BNT162b2 vaccine, an mRNA vaccine over 90% effective against clinical disease (*2,3*), and possibly against infection (*4*). As of March 29^th^ 2021, 51% of the population had received two doses (*5*). The impact of vaccination on SARS-CoV-2 transmission is now clearly seen, with a gradual decrease of the basic reproduction ratio (R_0_), which stood at 0.5 on March 19^th^ (*5*) despite a gradual easing of social distancing measures since the beginning of March 2021.

In Israel, individuals previously infected with SARS-CoV-2 were not eligible for vaccination until early March 2021. The duration of immunity following natural infection remains unknown but evidence shows that despite rapid waning of anti-spike SARS-CoV-2 IgG antibodies following natural infection (*6*), memory B cells persist at least 8 months (*7*). Vaccinating previously infected individuals with a single dose of BNT162b2 vaccine generates a boost-type IgG response up to 10 months post-infection (*8*). As a result, some countries including Israel now recommend a single dose of vaccine in infected individuals (*9*). The benefit of the second dose in such individuals remains uncertain.

Vaccine efficacy from a single dose of the BNT162b2 vaccine in uninfected individuals begins 14 days post-vaccination and reaches 50-80% between days 15-35) (*2*). Therefore, a proportion of patients will be infected after vaccination, mostly within the first 14 days post-vaccination. In Israel these individuals are not offered a second dose, based on the assumption that one dose confers immunity in infected individuals. However, immunogenicity in patients infected after one dose of vaccine has not been evaluated.

Our aim was to measure Anti-spike IgG response in a cohort of vaccinated healthcare workers according to SARS-CoV2 infection status (never infected or infected before or after vaccination) in order to inform vaccination policy.

## Methods

All employees of Ziv Medical Centre (ZMC), a hospital in Northern Israel, were offered vaccination with the BNT162b2 vaccine from December 2020. Before vaccination, Nucleocapsid (N) IgG antibody levels were measured in consenting workers, using a highly sensitive and specific SARS-CoV-2 IgG qualitative assay (Abbott, Abbot Park, US) (*10*), followed by a quantitative LIAISON SARS-CoV-2 S1/S2 IgG PCR test (DiaSorin, Saluggia, Italy) for verification purposes (*10*). Workers were also asked about evidence of previous infection with SARS-CoV-2. All the individuals with detectable IgG antibodies at baseline and/or evidence of a previous positive PCR test SARS-CoV-2 were considered previously infected.

AntiSARS-CoV-2 spike IgG levels were measured twice: around 21 days (range 15-35 days) and around 51 days (range 41-65 days) post-dose one, using the LIAISON Diasorin SARS-CoV-2 S1/S2 IgG assay (*10*).

All workers who developed COVID19-compatible symptoms during the study period (December 2020-March 2021) were PCR tested. Individuals with a positive PCR test were classified as infected post-vaccination.

Antibody levels were reported using geometric mean concentration (GMC) alongside 95% confidence intervals (95% CI). In the Israeli context, gender, ethnicity and time elapsed between infection and vaccination were not significantly associated with IgG levels (*8*) and were therefore not adjust for. Anti-SARS-CoV2 spike IgG antibody levels were compared among groups of ZMC workers according to the number of doses received (1 or 2) and their infection status (uninfected, infected prior to vaccination, infected after vaccination).The study was approved by Ziv Medical Center’s ethics committee (approval number: 0133–20-ZIV).

## Results

Of approximately 1,500 employees, 725 received at least one dose of vaccine and were tested at least once post-vaccination. Of these, 25 had evidence of pre-vaccination infection and 35 had evidence of infection post-dose 1 (Table), of which 32 before they were eligible for dose 2. These 32 individuals did not receive dose 2 as they were no longer eligible owing to their infected status. Not all these patients were tested twice (table).

**Table.**
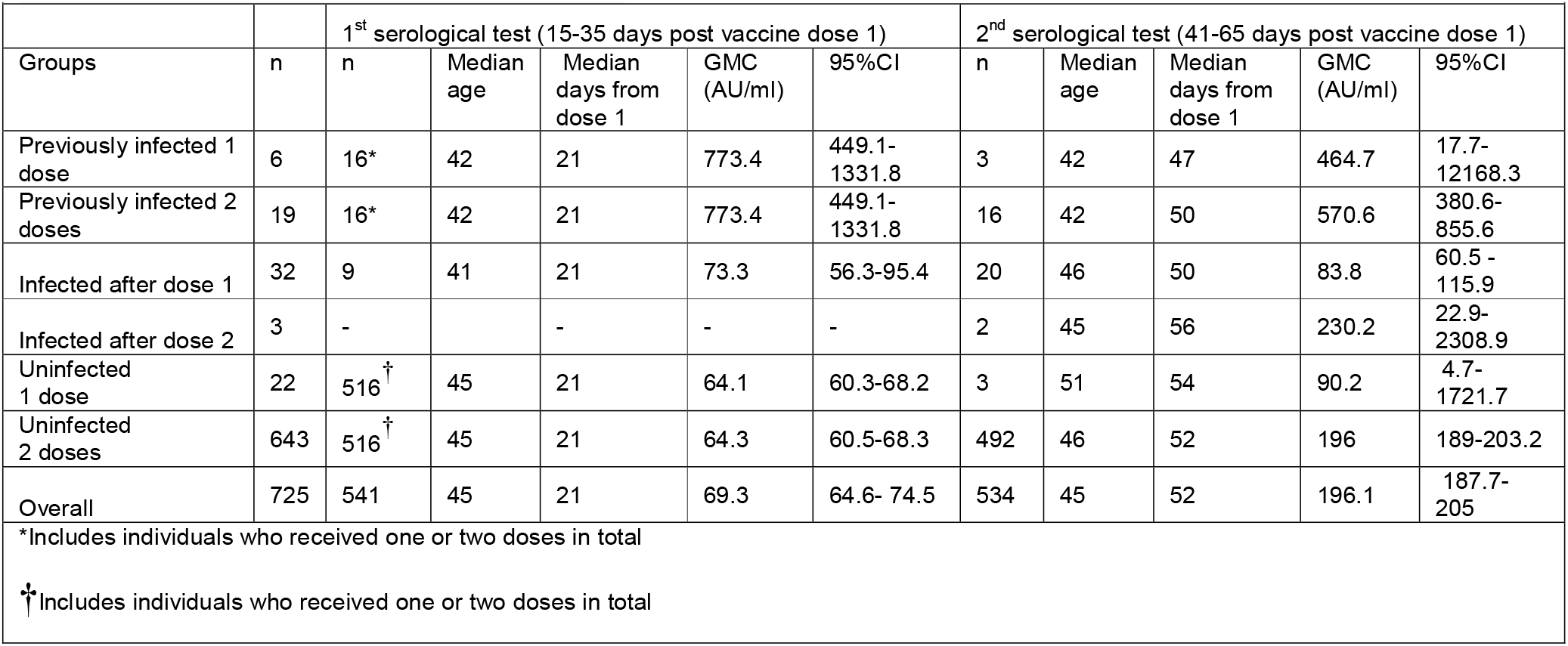
Geometric mean concentrations of anti-SARS-CoV-2 spike IgG antibodies among healthcare workers who received the BNT162b2 mRNA COVID-19 vaccine, Israel, December 2020 to March 2021

Compared with uninfected individuals, previously infected individuals who received one vaccine dose had high IgG titers at 21 days post-vaccine that remained high approximately 50 days post-vaccination (table), more than twice higher than previously uninfected individuals who received two doses (GMC 464.7 vs 196.1, table). Among those previously infected who received a second dose, IgG titers 50 days post-dose 1 (approximately 30 days post dose 2) had decreased compared with pre-dose 2 (GMC 570.6 vs 773.4, table) and was not significantly higher than those who received a single dose (table).

Among the 32 patients infected post-dose 1 and before being eligible for dose 2, 9 were tested around 21 days post vaccination, of which 8 were infected within 14 days of being vaccination and one between days 14-21. Among the 9, IgG levels at 21 days were comparable to those never infected (GMC 73.3 vs 64.3, table) and one order of magnitude lower than those infected prior to vaccination (GMC 73.3 vs 773.4, table). At 50 days post vaccination, IgG titers among the 20 patients infected post-dose 1 tested at this time point was 83.8 (95%CI 60.5-115.9), comparable to those previously uninfected who received a single dose, and lower than either those uninfected who received two doses or those infected prior to vaccination (table). Among the small number (n=3) of patients infected post-dose 2, IgG titers were comparable to those never infected (GMC 230.2 vs 196, table)

## Conclusions

Individuals infected with SARS-CoV-2 pre-vaccination have a strong and persisting response after one BNT162b2 dose. Dose 2 in those individuals has no impact on IgG titers. Several countries, including France and Israel recommend one vaccine dose for these individuals. Our data supports this policy.

Our study shows that when it comes to vaccinating infected individuals, the sequence of events matters. Individuals infected post-vaccination had IgG titers at 21 and 50 days similar to those never infected who received the same number of doses and much lower than those infected pre-vaccination. Individuals infected after a single dose of the Pfizer vaccine should therefore remain eligible for a second dose to ensure adequate protection levels. This implies a change in vaccination policy in many countries including Israel.

Our study presents limitations. First, our numbers are small. Notably, most previously infected who received one dose were not tested again at 50 days. Very few patients were infected after receiving their second dose, a finding compatible with the high effectiveness of the vaccine (*3*). Second, post-vaccination testing was symptom-based, and we could not detect asymptomatic patients infected post-vaccination. Our study focuses on the BNT162b2 vaccine and it is possible that these findings are not relevant to other vaccines. Lastly, our study measures circulating antibody levels and does not measure other components of the immune system. Interpreting the clinical significance of difference in IgG levels in the absence of correlates of protection remains a challenge. Nevertheless, this study demonstrates no benefit in receiving a second dose among individuals infected prior to vaccination, and more importantly, that infection after vaccination (dose 1 or dose 2) seems to have very little effect on immunogenicity. It is therefore important to not assume that individuals infected after their first dose are fully immune-these individuals need to receive a second dose of COVID-19 vaccine.

## Data Availability

The dataset used in the study contains confidential information and is not publicly available. The authors can be contacted to discuss requests for an anonymised version of the dataset.

## Funding

no external funding was received

## Conflct of interest

the authors declare no conflict of interest

## Biographical Sketch

Dr Kamal Abu Jabal is an internal medicine specialist since 2007 and was the senior physician at Ziv Medical Centre’s internal medicine department until September 2020. He is the head of Ziv Medical Centre’s COVID19 department since September 2020. His main research interests are liver disease and COVID19.

## Notes

### Competing Interest Statement

The authors have declared no competing interest.

### Author Declarations

The study was approved by ZMC ethics committee (013320ZIV).

## References

1. Our world in data Coronavirus (COVID-19) Vaccinations - Statistics and Research - Our World in Data

2. Polack FP, Thomas SJ, Kitchin N, Absalon J, Gurtman A, Lockhart S et al. Safety and efficacy of the BNT162b2 mRNA Covid-19 vaccine. N Engl J Med. 2020;383(27):2603–15. https://doi.org/10.1056/NEJMoa2034577 PMID: 33301246

3. Dagan N, Barda N, Kepten E, Miron O, Perchik S, Katz M, et al. BNT162b2 mRNA Covid-19 Vaccine in a Nationwide Mass Vaccination Setting. N Engl J Med. 2021 Feb 24;NEJMoa2101765. doi:10.1056/NEJMoa2101765

4. Thompson M, Burgess J, Naleway A, Tyner H, Yoon S, Meece J et al. Interim Estimates of Vaccine Effectiveness of BNT162b2 and mRNA-1273 COVID-19 Vaccines in Preventing SARS-CoV-2 Infection Among Health Care Personnel, First Responders, and Other Essential and Frontline Workers — Eight U.S. Locations, December 2020–March 2021. MMWR. March 29, 2021 / 70

5. Ministry of Health of Israel. [Coronavirus in Israel – general situation]. Jerusalem: Ministry of Health. [Accessed: 31 Mar 2021]. Hebrew. Available from: https://datadashboard.health.gov.il/COVID-19/general

6. Ibarrondo F, Fulcher J, Goodman-Meza D, Elliott J, Hofmann C, Hausner M et al. Rapid Decay of Anti-SARS-CoV-2 Antibodies in Persons with Mild Covid-19. N Engl J Med. 2020 Sep 10;383(11):1085–1087

7. Hartley GE, Edwards ESJ, Aui PM, Varese N, Stojanovic S, McMahon J et al. Rapid generation of durable B cell memory to SARS-CoV-2 spike and nucleocapsid proteins in COVID-19 and convalescence.Sci Immunol. 2020 Dec 22;5(54):eabf8891. doi:10.1126/sciimmunol.abf8891.

8. Abu Jabal K, Ben-Amram H, Beiruti K, Batheesh Y, Sussan C, Zarka S et al. Impact of age, ethnicity, sex and prior infection status on immunogenicity following a single dose of the BNT162b2 mRNA COVID-19 vaccine: real-world evidence from healthcare workers, Israel, December 2020 to January 2021.Euro Surveill. 2021 Feb;26(6):2100096. doi:10.2807/1560-7917.ES.2021.26.6.2100096.

9. French Health Authority. [Vaccination strategie against SARS-CoV-2-vaccination of individuals with a history of COVID-19 infection]. Paris: Ministry of Health. [Accessed 31 Mar 2021]. French. Available from: Haute Autorité de Santé - Stratégie de vaccination contre le SARS-CoV-2 - Vaccination des personnes ayant un antécédent de Covid-19 (has-sante.fr)

10. Turbett SE, Anahtar M, Dighe AS, Garcia Beltran W, Miller T, Scott H, et al. Evaluation of Three Commercial SARS-CoV-2 Serologic Assays and Their Performance in Two-Test Algorithms. J Clin Microbiol. 2020;59(1):59. https://doi.org/10.1128/JCM.01892-20 PMID: 33020186

